# General practitioner wellbeing during the COVID-19 pandemic: a qualitative interview study

**DOI:** 10.1101/2022.01.26.22269874

**Authors:** Laura Jefferson, Claire Heathcote, Karen Bloor

## Abstract

**Background:** Workload pressures and poor job satisfaction have been reported by UK general practitioners (GPs) for some time. The COVID-19 pandemic presented new challenges, with growing international evidence of its negative impact on GPs’ mental health and wellbeing. While there has been wide commentary on this topic, UK research evidence is lacking. Developing greater understanding of these lived experiences and subgroup differences is important as doctor wellbeing may affect the sustainability of health care systems and quality of patient care.

**Objectives:** To explore the lived experience of UK GPs during COVID-19, and the pandemic’s impact on their psychological wellbeing.

**Design and Setting:** In-depth qualitative interviews, conducted remotely by telephone or video call, with NHS GPs.

**Participants:** GPs were sampled purposively across three career stages (early career, established and late career or retired GPs) with variation in other key demographics. A comprehensive recruitment strategy used multiple channels. Data were analysed thematically using Framework Analysis.

**Results:** We interviewed 40 GPs; most described generally negative sentiment and many displayed signs of psychological distress and burnout. Causes of stress and anxiety related to personal risk, workload, practice changes, public perceptions and leadership, teamworking and wider collaboration and personal challenges. GPs described facilitators of their wellbeing, including sources of support and plans to reduce clinical hours or change career path.

**Conclusions:** A range of factors detrimentally affected the wellbeing of GPs during the pandemic and we highlight the potential impact of this on workforce retention and quality of care. As the pandemic progresses and general practice faces continued challenges, urgent policy measures are now needed.

**Strengths and limitations of this study:**

- While there is growing international evidence base demonstrating the impact of the COVID-19 pandemic on GPs’ wellbeing and much UK media coverage, this qualitative interview study provides much-needed research evidence of UK GPs’ lived experiences and wellbeing during COVID-19.
- 40 GPs were sampled purposively to include GPs with different demographic and practice characteristics.
- While there are no easy solutions to the problems highlighted, this research provides increased contextualised understanding of how these experiences may impact future workforce retention and the sustainability of health systems longer-term.
- Sub-group differences by gender and age are reported; highlighting a potential need for further research and support targeted at specific groups.
- Findings are necessarily limited to the time of data collection (Spring/Summer 2021); further tensions in general practice have since arisen, particularly regarding negative and misleading media portrayal.

## Introduction

Before the COVID-19 pandemic, rising demands on UK NHS general practitioners (GPs), including increasing work complexity and intensity and falling numbers of doctors, was leading to a growing gap between GP demand and supply.^1^ 80% of all doctors participating in a BMA survey appear to be at high or very high risk of burnout,^2^ with research suggesting primary care doctors are at highest risk.^3, 4^ Not only does chronic stress and burnout threaten the mental health of GPs, but it also presents challenges for the sustainability of the health care system and the quality of patient care. Pre-COVID-19, one in three GPs planned to leave medicine within five years^5^ and a shortage of 2,500 GPs was estimated to increase to 7,000 within five years if trends continued.^1^ The link between doctor wellbeing and patient safety has been demonstrated in a systematic review,^6^ while in general practice specifically, lower wellbeing has been associated with increased likelihood of reporting ‘near miss’ events and worse perceptions of patient safety.^7^

Clear new risks to workforce wellbeing occurred during the pandemic: GPs have experienced rapid change, risks of infection, remote working and reductions in face-to-face patient care. A growing international research evidence base has explored the impact of the pandemic on healthcare workforce wellbeing.^8-14^ Indeed, 31 studies in general practice were included in a recent systematic review of international literature.^15^ While these studies highlight pressures during the pandemic and impact on GPs’ psychological wellbeing, just three research studies including UK GPs were identified. One of these studies explores experiences of GPs with long-COVID, one focuses on one geographical location, and one presents the findings of UK GPs alongside other countries.

We sought to address this evidence gap, by exploring the lived experience of UK GPs during COVID-19, and the pandemic’s impact on their psychological wellbeing.

## Method

We adopted an exploratory qualitative methodology, conducting qualitative interviews to understand UK GPs’ lived experiences and wellbeing during COVID-19. While our analytical approach was inductive in nature and a pre-defined theoretical framework was not imposed, our approach was guided by our existing knowledge of relevant literature. We interpret our findings within the policy context using the ABC of doctors’ needs,^16^ which is based on Deci and Ryan’s self-determination theory.^17^

A multidisciplinary team developed and piloted topic guides in consultation with an expert panel comprising several GPs and a project steering committee consisting of international experts in organisational psychology, NHS mental health and senior Royal College of General Practitioner (RCGP) representatives. Three patient representatives were also involved in the design and implementation of this research. Interviews were semi-structured in nature, using topic guides to explore GPs’ wellbeing during the pandemic, encouraging reflections on their working lives and wellbeing before the pandemic, views around challenges during the pandemic, facilitators of improved working practices, future intentions, motivations and thoughts on how to improve GPs’ working lives.

### Sampling and recruitment

We sampled GPs purposively across three career stages: ‘early-career GPs’ (in final stages of training and first five years of practice); ‘established GPs’ and ‘late-career GPs (including retired GPs returning to practice during COVID-19). We sampled for variation in key demographics including ethnicity, age, gender, contract type and local area characteristics (geographical spread, deprivation level and COVID-19 rates) using a comprehensive, multi-channel recruitment strategy. We received a good response through social media dissemination, but to ensure variety and reduce potential bias we also recruited through our regional deanery, local and national networks, respondents to the GP Work Life Survey and the RCGP late-career and recently retired group.

Potential participants were asked to complete a brief survey to provide contact details and basic demographic information, and sent Participant Information Leaflets and Consent Forms. GPs meeting the sampling framework were contacted to arrange virtual interviews, conducted via zoom or telephone. To thank participants for their time, we provided a £100 payment.

### Analysis

We used transcriptions and recordings to analyse data thematically, facilitated using NVivo 12 data sorting software (QSR International Pty Ltd, 2018). Our approach to analysis was inductive, with themes emerging from the data rather than using pre-specified theory. We used Framework Analysis^18^ following the steps described in Table 1. Two researchers (LJ and CH) coded the interviews independently, checking a 20% sample for consistency. Qualitative researchers met weekly to enable triangulation; refining the coding framework as analysis progressed. No member checking was needed.

**Table 1:** Process of Framework Analysis

| Stage of Analysis | Description |
| --- | --- |
| Managing the data | We managed transcriptions using Nvivo 12 software (QSR International Pty Ltd, 2018) to supplement the researchers' analytical thinking and familiarisation with the data. |
| Familiarisation | Both researchers (CH and LJ) that undertook interviews immersed themselves in the data by reading and re-reading transcripts, listening to audio recordings and producing detailed notes for each interview in order to help facilitate the following analysis stages. |
| Identifying a thematic framework | Researchers independently developed two thematic frameworks and met on multiple occasions to discuss and refine these into one thematic framework. This was tested on 4 transcripts prior to use, and further iterations continued to be made through discussion with the study team as the coding developed. |
| Indexing the data | Both researchers then indexed, or coded the interview data according to 10 themes and 95 subthemes which were identified in the thematic framework. Data were re-coded where needed whenever revisions to the coding framework were made. |
| Charting | Once coding was complete, we explored the relationships between themes using mindmaps, research team discussions and creation of overarching themes, or 'supercodes.' This process identified six overarching themes made up of 30 subthemes. We also explored categories of participants, particularly focusing on relationships between career stage, gender, job role, ethnicity, previous or current experience of mental illness and working in a deprived geographical area. We used count data to explore potential trends in analysis, though this did not replace in-depth qualitative analysis of the data, which was facilitated through mapping themes according to these key characteristics. |
| Mapping and interpretation | In order to go beyond the purely descriptive account of the data and develop wider meanings about links between phenomena and subgroups of participants, we mapped themes to build patterns in the data, bearing in mind the original research objectives and also exploring negative or deviant cases to explore alternative explanations for the data. |

### Reflexivity

We maintained a reflexive approach throughout the design and analysis stages to limit potential for preconceptions to influence research findings. All researchers were female, with non-medical backgrounds. We undertook researcher triangulation (during data collection and analysis) and discussed findings with a committee of experts, GPs and patients.

## Results

### Sample characteristics

Interviews with 40 GPs took place between March and June 2021, lasting between 43 and 72 minutes. Participants were from a range of career stages: 13 ‘early career’, 19 ‘established’ and 8 ‘late-career’ or retired GPs. This is reflected in the spread of ages detailed in Table 2. Twenty GPs were aged 30-39, and we interviewed more women than men (29/40). There was a slightly higher proportion of white GPs in our sample to those reported nationally (67.5% compared to 56.6% nationally^19^). We interviewed more salaried GPs (17) than other job roles, followed by GP partners (14). GPs in our sample worked between 1 and 8 clinical sessions per week (median 6, interquartile range 3.63) and almost half of participants (n=18) also held additional roles alongside their clinical workload (e.g. practice management, teaching, research, mentoring, national or local leadership roles). Six were working as locum GPs or undertook additional locum work. Four GPs reported having had a confirmed COVID-19 diagnosis and a further eight suspected having had COVID-19 when testing was not available at the start of the pandemic. 10 participants were working in areas of high deprivation, nine in areas with pockets of deprivation, four worked in rural or semi-rural locations and four described serving a large elderly population.

**Table 2:** Participant characteristics

| Career stage | N | (%) |
| --- | --- | --- |
| *Early* | 13 | 32.5 |
| *Established* | 19 | 47.5 |
| *Late* | 8 | 20.0 |
| Gender |  |  |
| *Male* | 11 | 27.5 |
| *Female* | 29 | 72.5 |
| Age |  |  |
| *< 30* | 3 | 7.5 |
| *30 - 39* | 20 | 50.0 |
| *40 - 49* | 9 | 22.5 |
| *50 - 59* | 6 | 15.0 |
| *>60* | 2 | 5.0 |
| Ethnicity |  |  |
| *Ethnic minority Groups* | 10 | 25.0 |
| *White British* | 27 | 67.5 |
| *White non-British* | 3 | 7.5 |
| Location |  |  |
| *England - East* | 3 | 7.5 |
| *England - London* | 5 | 12.5 |
| *England - North East* | 1 | 2.5 |
| *England - North West* | 3 | 7.5 |
| *England - South East* | 3 | 7.5 |
| *England - South West* | 4 | 10.0 |
| *England - West Midlands* | 5 | 12.5 |
| *England - Yorkshire and Humber* | 14 | 35.0 |
| *Northern Ireland* | 2 | 5.0 |
| Job role |  |  |
| *GP trainee* | 6 | 15.0 |
| *GP retainer* | 1 | 2.5 |
| *Salaried GP* | 17 | 42.5 |
| *GP partner* | 14 | 35.0 |
| *Retired GP* | 2 | 5.0 |
| Clinical sessions |  |  |
| *Median (IQR)* | 6 (3.63) |  |
| *1-4* | 11 | 27.5 |
| *5-7* | 16 | 40.0 |
| *≥8* | 9 | 22.5 |
| *Retired* | 2 | 5.0 |
| *Unknown* | 2 | 5.0 |
| *Portfolio roles* | 18 | 45.0 |
| Area demographics |  |  |
| *Highly deprived* | 10 | 25.0 |
| *Pockets of deprivation* | 9 | 22.5 |
| *Rural or semi-rural* | 4 | 10.0 |
| *Large elderly population* | 4 | 10.0 |
| COVID history |  |  |
| *Suspected COVID* | 8 | 20.0 |
| *COVID diagnosis* | 4 | 10.0 |

**Table 3:** Participant characteristics and descriptive IDs.

| Descriptive ID | Career stage | Age | Ethnicity | Gender | Role | Region |
| --- | --- | --- | --- | --- | --- | --- |
| GP1,MpartnerEst | Established | 30 - 39 | White British | Male | GP partner | England - Yorkshire & Humber |
| GP2,FsalariedEst | Established | 30 - 39 | White British | Female | Salaried GP | England - Yorkshire & Humber |
| GP3,FpartnerLate | Late | 50 - 59 | White British | Female | GP partner | England - North East |
| GP4,MsalariedEarly | Early | 30 - 39 | White British | Male | Salaried GP | England - Yorkshire & Humber |
| GP5,FsalariedEst | Established | 40 - 49 | White British | Female | Salaried GP | England - North West |
| GP6,FsalariedEarly | Early | 30 - 39 | Asian British | Female | Salaried GP | England - Yorkshire & Humber |
| GP7,MpartnerLate | Late | 50 - 59 | Asian / Asian British - Pakistani | Male | GP partner | England - Yorkshire & Humber |
| GP8,FsalariedEarly | Early | 30 - 39 | White British | Female | Salaried GP | England - Yorkshire & Humber |
| GP9,FpartnerEst | Established | 40 - 49 | White British | Female | GP partner | England - Yorkshire & Humber |
| GP10,FtraineeEarly | Early | < 30 | Asian / Asian British - Indian | Female | GP trainee | England - London |
| GP11,FsalariedEst | Established | 30 - 39 | Asian / Asian British - Pakistani | Female | Salaried GP | England - West Midlands |
| GP12,MtraineeEarly | Early | < 30 | White British | Male | GP trainee | Northern Ireland |
| GP13,FtraineeEarly | Early | 30 - 39 | White - Irish | Female | GP trainee | Northern Ireland |
| GP14MpartnerEst | Established | 30 - 39 | Asian / Asian British - Indian | Male | GP partner | England - West Midlands |
| GP15,FretainerEarly | Early | 30 - 39 | White British | Female | GP retainer | England - Yorkshire & Humber |
| GP16,MsalariedEarly | Early | 30 - 39 | White British | Male | Salaried GP | England - West Midlands |
| GP17,Mretired | Late | >60 | White British | Male | Retired GP partner | England - South East |
| GP18,FpartnerLate | Late | 50 - 59 | White - Other | Female | GP partner | England - North West |
| GP19,FpartnerLate | Late | 50 - 59 | White British | Female | GP partner | England - South West |
| GP20,FtraineeEarly | Early | 30 - 39 | White British | Female | GP trainee | England - Yorkshire & Humber |
| GP21,FsalariedEst | Established | 40 - 49 | Other ethnic group - Arab | Female | Salaried GP | England - West Midlands |
| GP22,FsalariedEst | Established | 30 - 39 | White British | Female | Salaried GP | England - South West |
| GP23,FsalariedLate | Late | 50 - 59 | White British | Female | Salaried GP | England - East |
| GP24,FpartnerLate | Late | 50 - 59 | White British | Female | GP partner | England - South West |
| GP25,FsalariedEst | Established | 40 - 49 | Asian / Asian British - Indian | Female | Salaried GP | England - London |
| GP26,FtraineeEarly | Early | 30 - 39 | White British | Female | GP trainee | England - Yorkshire & Humber |
| GP27,MpartnerEst | Established | 40 - 49 | White British | Male | GP partner | England - Yorkshire & Humber |
| GP28,MsalariedEarly | Early | 30 - 39 | White British | Male | Salaried GP | England - London |
| GP29,FtraineeEarly | Early | 30 - 39 | Asian | Female | GP trainee | England - East |
| GP30,FpartnerEst | Established | 40 - 49 | White British | Female | GP partner | England - North West |
| GP31,FsalariedEst | Established | 40 - 49 | White British | Female | Salaried GP | England - South East |
| GP32,MpartnerEst | Established | 40 - 49 | White British | Male | GP partner | England - West Midlands |
| GP33,FsalariedEst | Established | 30 - 39 | Black - African | Female | Salaried GP | England - London |
| GP34,FsalariedEst | Established | 30 - 39 | White British | Female | Salaried GP | England - South West |
| GP35,FpartnerEst | Established | 30 - 39 | White British | Female | GP partner | England - East |
| GP36,FpartnerEst | Established | 30 - 39 | White British | Female | GP partner | England - Yorkshire & Humber |
| GP37,MpartnerEst | Established | 40 - 49 | White British | Male | GP partner | England - Yorkshire & Humber |
| GP38,FsalariedEst | Established | 30 - 39 | Asian / Asian British - Pakistani | Female | Salaried GP | England - South East |
| GP39,FsalariedEarly | Early | <30 | White British | Female | Salaried GP | England - Yorkshire & Humber |
| GP40,Fretired | Late | >60 | White - Other | Female | Retired GP partner | England - London |

### Thematic findings

Overarching themes highlighting 1) the impact of the pandemic on GPs’ psychological wellbeing, 2) causes of stress and anxiety and 3) facilitators that improved GPs’ working lives are described. These are displayed graphically in Figure 1.

**Figure 1:**
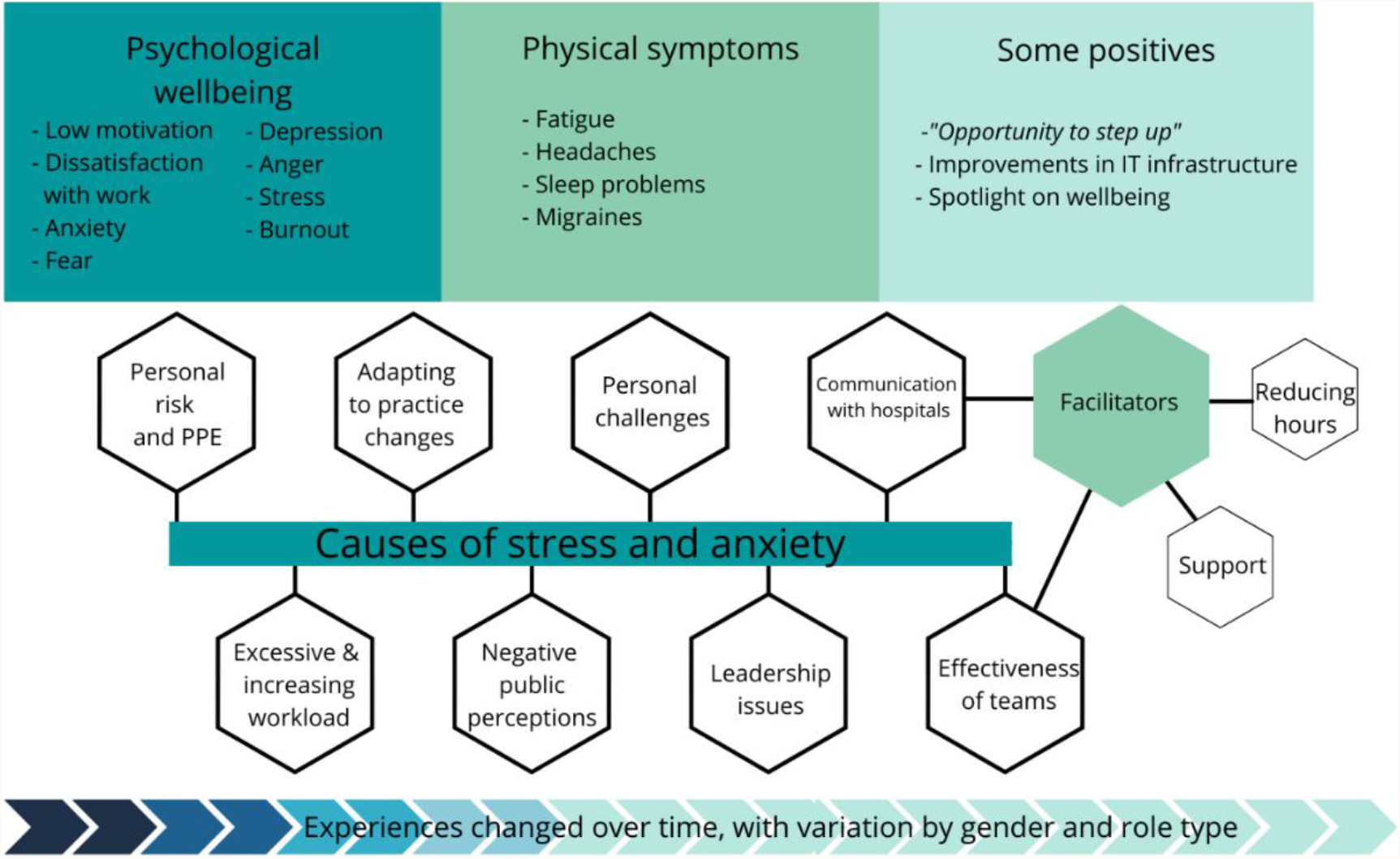
Graphical representation of the study findings.

### Psychological wellbeing

GPs talked about low motivation, dissatisfaction with work, frustration and anger during interviews, which they described as having been particularly difficult during the winter of 2020. For some this related to general stress of the pandemic (social isolation, lack of enjoyment in things and pressures of home-schooling). Work-related feelings of stress and anxiety were, however, very widely expressed. Often referred to as being overwhelmed, GPs described their work as *“all consuming”* (GP2,FsalariedEst) and having a “*background level of anxiety”* (GP3,FpartnerLate).

Causes of stress and anxiety altered during the course of the pandemic. At the start of the pandemic many commented on concerns around managing adaptations to work (e.g. movement to remote working and development of hot sites), but also dealing with uncertainty around what lay ahead. GPs described fear of the unknown and potential risk to themselves and their families. Anxiety increased as levels of unmet patient need grew from the autumn of 2020 onwards; there were concerns about future demand, as well as support available for patients’ mental and physical needs.

Five GPs reported having clinically diagnosed mental health problems; all were female (though with variation in age and job roles). One GP described her experience, which displayed signs characteristic of burnout, and needing to take time off to recover:

> *“You’re just filling and filling the bucket, and at some point it will overspill. And you’ve just got to hope that you don’t miss something really important… So I want to remove myself from that situation for at least a period of time, just while I rebuild my armour I suppose and see if I want to do it again*.*” GP34,FsalariedEst*

Many GPs described the negative impact on their families and relationships, and held concerns about quality of patient care due to increasing impatience or fear of making mistakes due to extreme fatigue. Difficulties with sleep and fatigue were common. Three GPs (one of whom experienced long COVID) described difficulties with concentration, resulting in driving incidents.

#### Stigma and presenteeism

GPs tended to downplay experiences of stress and, despite the impact on their mental wellbeing, many did not seek formal support:

> *“I am normally very ‘just get on with it’ in life. I massively took a dive. Just very anxious, not in a way that I needed any kind of help… but just completely changed who I was. I was a bit of a mess, much like most of us were*.*” GP26,FtraineeEarly*

GPs described reluctance to seek support because of stigma and guilt from taking time off as this would burden their colleagues without a *“buffer in the system”* (GP3,FpartnerLate). All had worked additional clinical sessions to cover absences, which increased during the pandemic due to mental wellbeing or self-isolation of colleagues. This appeared more problematic for GP partners and smaller practices.

> *“I think we all were put under huge stress and people have gone off sick that have never been sick. And I think people have just cracked up basically, but the trouble is, it’s like a domino effect” GP24,FpartnerLate*

#### Positive emotions

Approximately half of participants (17/40) expressed some positive comments when reflecting on their wellbeing during the pandemic. Many of these related to their enjoyment of work and doing a job they loved. Four recently qualified GPs welcomed the challenge and ability to ‘step up’ during the pandemic.

### Causes of stress and anxiety

#### Personal risk

Most interviewees reported fear of putting themselves and family members at risk, particularly at the start of the pandemic. GPs in high risk categories (older GPs, GPs from minority ethnic groups or those with asthma) described particular concerns. For example, a GP from an ethnic minority group described:

> *“I didn’t feel I was particularly protected in any way, you know, they just expect you to get on with it” GP7,MpartnerLate*.

Changing guidance around implementation of ‘hot’ sites, use of and access to PPE heightened anxiety. GPs were frustrated and felt neglected compared to hospital colleagues due to lower standards of PPE, even in COVID-19 ‘hot sites.’

> *“The psychiatrists were being fitted with FFP3 masks, specialist masks… working at home doing telephone reviews, and us in primary care and our district nurses… going out to visit cancer people were given flimsy surgical masks and told that these will be fine, get on with it… we felt disappointed that we were neglected” GP30,FpartnerEst*

#### Workload

GPs described workload issues before COVID-19, with treatment advances and shifting care out of hospitals adding pressure. The vast majority of GPs felt their workload had increased during the pandemic, reducing their wellbeing further.

> *“It’s a different world, isn’t it? I mean I think I thought I was busy [before COVID], but I didn’t have a clue what busy was, basically. I just can’t believe the workload explosion since COVID*… *it was stressful [before COVID], but I had my head above water*.*” GP24,FpartnerLate*

Working 12-14 hour days and additional unpaid administration sessions were commonplace. Patient demand for urgent on-the-day appointments was described as unmanageable, and practices also struggled to meet ‘non-urgent’ demand within reasonable timeframes.

> *“So most days there were 50 or 60 contacts on that appointment list where the RCGP says that they reckon the safe limit is about 30. So probably double*.*” GP8,FsalariedEarly*

GP partners, in particular, commented on increases in administrative workload at the start of the pandemic; reading and implementing sometimes contradictory guidance from multiple sources which evolved daily. At the start of the pandemic, though, the increased management workload was balanced by initial reduced patient demand. Management workload increased again during the planning and implementation of the vaccination programme, with additional time pressures from cleaning and PPE measures.

GPs reflected that patient demand became most challenging from the end of summer 2020 onwards, particularly from late presentations with more serious pathologies; leading to greater workload and emotional strain. Higher demand from patients with mental health problems also increased workload, alongside difficulties in consulting these patients remotely and lack of support services:

> *“Our mental health service is shocking… mental health services play ping pong between themselves… IAPT say, oh, too severe for us, and the secondary care mental health service say, oh, no, not severe enough for us, we’re not dealing with that. And then they just fall into this black hole*.*” GP35,FpartnerEst*

#### Practice changes

Participants described the many changes that the pandemic had brought about, including new triage systems, use of remote consultations, the vaccination rollout and changes for trainees. Some associated these changes with stress and increased workload, but there was a general sentiment that the pandemic had provided a positive impetus for technological development. GPs described the importance of triage systems for prioritisation and reallocating patients during staff absences. E-consultation systems were perceived to increase demand due to greater accessibility:

> “*Now eConsults have come in there’s no barrier… there’ll be 200 eConsults on a Monday that we have to deal with as well as all the other general practice workload and the vaccination programme and PCNs, and it’s just really unsustainable and unsafe*.*” GP30,FpartnerEst*

There were mixed emotions around the movement to telephone and video consultations, which were viewed positively for minor conditions, reducing attendances and enabling more focused face-to-face appointments. GPs in multi-site practices covering large geographical areas described their increased ability to share workload across practices. GPs also described feeling isolated, ‘decision fatigue’ and felt that consultations lacked personal contact with patients, which had encouraged their career choice. While telephone consultations were well-received amongst younger and working patients, there were concerns around inequalities in access and potential missed diagnoses. These concerns were particularly expressed by trainee and early-career GPs.

### Vaccination rollout

The vaccination programme was described as a great morale booster, coming at a time when many GPs and the wider public needed hope. GPs described working additional hours to manage vaccinations, but with a sense of teamwork and pride.

> *“There was a point when we were doing the 80 year olds where you had to vaccinate 14 people to save one life. And I’m feeling tearful about it even now. Like just the actual practical difference that you could make in a terrible situation*.*” GP34,FsalariedEst*

Practices had also faced workload increases due to patient queries about vaccinations and GPs expressed frustration with public messaging around the vaccination rollout.

#### Public perceptions and leadership

Negative public perceptions of general practice greatly impacted GPs’ wellbeing and was one of the most widely cited causes of stress. Patients facing problems with access or referrals became increasingly frustrated, and GPs felt this was fuelled by negative media portrayals, described by participants as *“GP bashing*.*”* GPs described *“simmering discontent amongst communities”* (GP28,MsalariedEarly) who they felt had been *“whipped up to a frenzy by the government and by the media”* (GP24,FpartnerLate).

Sixteen GPs described positively the outpouring of appreciation for NHS workers at the beginning of the pandemic, but most felt that public appreciation was eroded due to inaccurate messaging from the government, NHS England and the media about general practice being closed:

> *“That was really upsetting at one point, thinking that people thought we were closed. I was like, I’ve been working my socks off, I’ve been working at COVID hubs or I’ve been doing back-to-back telephone consulting… no matter what we do or what we try, people just assume that we’re not working hard enough*.*” GP10,FtraineeEarly*

GPs expressed frustration around national decision-making, which they felt had directly risked NHS capacity and heightened anxiety in anticipation of repeat waves of the pandemic. Communication about delays in out-patient appointments and routine surgery was seen as vital, as were government campaigns encouraging health awareness about common illnesses and more signposting to appropriate specialists.

Retired GPs described lengthy bureaucratic processes prohibiting them from returning to practice; certain training requirements were viewed as unnecessary for remote working and one described the process taking two months. Two volunteered to support practices and vaccinations, but their offers were declined.

#### Wider collaboration

17/40 interviewees felt that the pandemic offered opportunities to foster collaboration across Primary Care Networks (PCNs), hospitals, community and wider services. A greater sense of camaraderie and improved working across PCNs was reported, with groups of practices ‘pulling together’ during the vaccine rollout.

A minority (2/40) reported greater access to specialist support from hospitals, with 12 GPs describing conflict between primary and secondary care. Lengthy hospital waiting lists and some service closures increased workload for GPs, who felt they were the only support for some high-risk patients:

> *“Eating disorder services stopped. They just stopped. So for a nine month period any new referrals, you couldn’t refer. And there wasn’t an alternative. So we set up a high risk list to look after the highest risk eating disorders patients. … Mental health services, closed to routine referrals. They would only see suicidal people*.*” GP34,FsalariedEst*

#### General practice teams

Experiences and perceptions of the effectiveness of practice teams varied, affecting GP wellbeing and ability to cope with challenges. Isolation from teams was problematic particularly for early-career GPs who lacked support and found it difficult to integrate. Concerns were raised around trainees’ wellbeing, feeling that they had been used *“as cannon fodder”* in frontline hospital roles and had faced much disruption to their training. Disproportionate numbers of women raised difficulties with teams (15/18 GPs).

30/40 GPs cited examples of good teamworking and described a sense of pulling together during the pandemic. An increased focus on personal and team mental wellbeing was reported, though some participants were disenchanted with initiatives that sought to improve ‘resilience’ as they felt that this placed the onus of responsibility at an individual rather than structural level. Others suggested wellbeing support was perhaps more easily adopted by larger practices with greater infrastructure. Team ‘huddles’ were used to debrief on complex cases, provide social support and share anxieties, but small rooms and safe distancing in some practices prohibited staff meetings. Shared breaks provided opportunities to raise difficulties informally, which was important to some who felt less inclined to seek formal support either due to workload pressures or stigma.

#### Personal challenges

Negative financial impacts of the pandemic were described by five GPs, mostly due to reduced availability of locum work, though one GP from a University practice described a reduction in practice earnings and associated stress due to reduced student/patient numbers. Challenges of home-schooling and reduced access to childcare were discussed by 14 GPs (12/14 were women), who described juggling telephone consultations and administrative work with childcare:

> *“So I was at home trying to get through more patients than normal remotely, trying to learn the technology and I had my children at home, so it was huge. I can remember feeling just running on adrenaline and just feeling constantly stressed*.*” GP30,FpartnerEst*

### Facilitators

#### Informal and formal support

Interviewees sought informal support through family and friends (28/40), colleagues (29/40) and peers (15/40). They described the benefits of talking to other medics who could relate to their experiences; this was particularly important to trainees, some of whom were isolated from family and other networks. There appeared to be good awareness of the different formal support structures available; ranging from coaching and mentoring support (used by 13/40) to more formal mental health support. Only two male participants discussed using these support services, and, similarly, gender differences were apparent in discussion of approaches to ‘self-care’; only three of 17 GPs discussing these techniques were male.

#### Reducing clinical hours and future plans

Eight GPs (6/8 women) discussed reducing their clinical sessions or developing portfolio careers in order to manage work pressure and support wellbeing. There was greater variation in the number of clinical sessions reported by women (median: 6, interquartile range: 3.0) than men (median 6, interquartile range 1.88) as some women had low numbers of clinical sessions, described as a reaction to risk of burnout and seeking work-life balance.

Portfolio careers (e.g. including teaching and mentoring) provided an opportunity to achieve greater balance, while others planned to specialise, become locums, work abroad or retire. GPs were concerned about retention, particularly of those approaching retirement. Greater use of retainer schemes or a phased retirement stage were seen as opportunities to reduce workload, stress and retain GPs.

## Discussion

### Summary

Our interviewees offered in-depth accounts of their experiences during the COVID-19 pandemic, highlighting an exacerbation of prior difficulties which, for some, had led to dissatisfaction with work and mental health problems. Some GPs planned to reduce their clinical or overall working hours, take on locum work, work abroad or retire. GPs described feelings characteristic of burnout and raised concerns around quality of patient care.

Pressures changed as the pandemic evolved. Early on, GPs experienced stress, rapid change, uncertainty and personal risks, but this time also catalysed technological change. Later, GPs faced anxiety relating to unmet patient need, delayed presentations and growing demand, particularly for mental health support, while negative patient perceptions and media portrayal of practices being ‘closed’ during this time increased GPs’ work stress and reduced job satisfaction. There were calls for improved public relations from leadership bodies in order to counteract inaccuracies in the media and to improve health literacy, particularly as uptake of e-consultation services was perceived as increasing patient demand.

A greater sense of camaraderie and working across PCNs was reported, particularly with vaccination delivery. Effective team-working was seen as vital and GPs welcomed an increasing focus on wellbeing. They also, however, described a culture of presenteeism; exacerbated during the pandemic due to staff absences and, for some, a sense of stigma around doctors’ mental health.

### Comparison with existing literature

While this research outlines key sources of stress for GPs that have been the subject of much recent commentary, to our knowledge this is the first reported qualitative study focused on UK GPs’ psychological wellbeing during the pandemic and this research also offers insights into potential subgroup variations. International literature highlights similar trends in GP wellbeing during the pandemic - doctors from varied settings report increased rates of burnout, related to high workload, job stress, time pressure and limited organisational support.^15,20^ International studies have found higher stress in general practice doctors compared to other healthcare workers and settings.^10, 21, 22^ The expanding public commentary and campaigns from UK doctor groups highlight the need to support the GP workforce.^23^

Subgroup variations in GPs’ experiences are important to understand as the pandemic progresses and workforce pressures continue. Our research revealed different effects on men and women GPs and different use of support services. This is consistent with international literature which reports gender differences in stress, burnout, anxiety and depression^9, 21, 22, 24-27^ and greater job strain amongst women in dual-doctor marriages during the pandemic.^28^ These differences may arise as a result of gendered social norms around willingness to disclose difficulties, or due to socially constructed gender roles in the home that proliferated during COVID-19 lockdowns, negatively impacting women in employment.^29, 30^ Our research also suggests gender differences may exist in GPs’ perceptions around effective teamworking; perhaps highlighting women’s differential support needs or expectations. Women may require targeted interventions to support their wellbeing and encourage continued participation, particularly as they were more likely to report future plans to reduce clinical sessions or adopt portfolio roles. GP partners may also require targeted support as they described greater pressures associated with management workload due to changes to service delivery, staff shortages and vaccination rollout, which supports other recent studies showing an association between older age and higher stress in GPs.^25, 31, 32^ Further research may be needed to explore recently qualified and trainee GPs’ experiences as our findings suggest they have faced differing challenges that may affect longer-term retention and wellbeing.

### Strength and limitations

This research provides rich and contextualised understanding of the experiences of a varied sample of GPs during the pandemic, which our recent systematic review (currently under review) identified as lacking from a UK setting. While there may be selection bias in the views expressed by GPs willing to share experiences, our interview findings are consistent with other international research and wider commentary on this topic. Our findings are necessarily limited to the time of data collection (Spring/Summer 2021); further tensions in general practice have since arisen, particularly regarding negative and misleading media portrayal.^33^

### Implications for research, policy and practice

This research demonstrates the effect of the pandemic on GP wellbeing, with potential wider impacts, for example around workforce retention and patient safety; highlighting a need for national and local intervention. Using Deci and Ryan’s self-determination theory^17^, a recent GMC report^16^ described the *“ABC of doctors’ needs”*, advising that doctors’ sense of autonomy, belonging and competence need to be promoted for them to thrive in their working lives. All three components have been threatened during the pandemic. GPs’ ability to control and influence their work has reduced, and patient frustrations and media blaming of GPs has affected their sense of belonging and competence. There is a need for policy to support GPs, prevent work stress and foster collaborations across wider teams.

Further research could explore these findings more widely through quantitative methods, preferably with some comparison with pre-pandemic wellbeing scores. E-consultation systems, which appear to have increased demand, could be further evaluated, as should planned schemes to supplement the GP workforce with other non-medical staff through the Additional Roles Reimbursement Scheme that formed part of recent GP contract revisions.^34^

## Conclusion

The COVID-19 pandemic created some positive impacts on general practice - changing working systems, increasing wider team-working and placing a spotlight on staff wellbeing. Nevertheless, a range of factors affected the wellbeing of GPs detrimentally during the pandemic, and substantial challenges to GPs remain. This could affect workforce retention, quality of care and the sustainability of health systems longer-term. Targeted support strategies may be required to address the subgroup variations, particularly the apparently more detrimental effects on women and on early-career GPs.

## Data Availability

All data produced in the present study are available upon reasonable request to the authors

## Acknowledgements

We are grateful to the GPs that gave their time and support through participating in this research. We would like to thank the members of our Project Steering Committee meeting for their contributions throughout the design and conduct of this study: Prof Dame Clare Gerada, Prof Michael West CBE, Prof Michael Holmes and our Patient and Public Representatives for their contributions throughout: Patricia Thornton, Stephen Rogers and Emma Williams. We also thank Dr Shanthi Antill and Drs Madeleine and Drew Bradman for their contributions during the design and development of this study, and Prof Katherine Checkland and Prof Matt Sutton for supporting the project and enabling recruitment of participants through the GP Worklife Survey.

## References

1. King’s Fund. Closing the gap report. Chapter 7: modelling the impact of reform and funding on nursing and GP shortages: Available online: https://www.kingsfund.org.uk/sites/default/files/2019-03/closing-the-gap-health-care-workforce-full-report.pdf#page=107 (last accessed 05/08/21). 2019.

2. BMA. Caring for the mental health of the medical workforce. Available online: https://www.bma.org.uk/media/1365/bma-caring-for-the-mental-health-survey-oct-2019.pdf (last accessed 05/08/21). London 2019.

3. GMC. The state of medical education and practice in the UK. Available online: https://www.gmc-uk.org/about/what-we-do-and-why/data-and-research/the-state-of-medical-education-and-practice-in-the-uk (last accessed 05/08/21). 2019.

4. McKinley N, McCain RS, Convie L, Clarke M, Dempster M, Campbell WJ, et al. Resilience, burnout and coping mechanisms in UK doctors: a cross-sectional study. BMJ Open. 2020;10(1):e031765.

5. Gibson J, Sutton M, Spooner S, Checkland K. Ninth National GP Worklife Survey.. University of Manchester: Policy Research Unit in Commissioning and the Healthcare System Manchester Centre for Health Economics. 2018.

6. Hall LH, Johnson J, Watt I, Tsipa A, O’Connor DB. Healthcare Staff Wellbeing, Burnout, and Patient Safety: A Systematic Review. PLoS One. 2016;11(7):e0159015.

7. Hall LH, Johnson J, Watt I, O’Connor DB. Association of GP wellbeing and burnout with patient safety in UK primary care: a cross-sectional survey. Br J Gen Pract. 2019;69(684):e507–e14.

8. Di Monte C, Monaco S, Mariani R, Di Trani M. From Resilience to Burnout: Psychological Features of Italian General Practitioners During COVID-19 Emergency. Front Psychol. 2020;32:567201.

9. Dutour M, Kirchhoff A, Janssen C, Meleze S, Chevalier H, Levy-Amon S, et al. Family medicine practitioners’ stress during the COVID-19 pandemic: a cross-sectional survey. BMC Fam Pract. 2021;22(1):36.

10. Rossi R, Socci V, Pacitti F, Mensi S, Di Marco A, Siracusano A, et al. Mental Health Outcomes Among Healthcare Workers and the General Population During the COVID-19 in Italy. Front Psychol. 2020;32:608986.

11. Sitanggang FP, Wirawan GBS, Wirawan IMA, Lesmana CBJ, Januraga PP. Determinants of Mental Health and Practice Behaviors of General Practitioners During COVID-19 Pandemic in Bali, Indonesia: A Cross-sectional Study. Risk Management & Healthcare Policy. 2021;32:2055–64.

12. Sotomayor-Castillo C, Nahidi S, Li C, Hespe C, Burns PL, Shaban RZ. General practitioners’ knowledge, preparedness, and experiences of managing COVID-19 in Australia. Infection, Disease and Health. 2021.

13. Ta SB, Ozceylan G, Ozturk GZ, Toprak D. Evaluation of Job Strain of Family Physicians in COVID-19 Pandemic Period-An Example from Turkey. J Community Health. 2020.

14. Trivedi N, Trivedi V, Moorthy A, Trivedi H. Recovery, restoration, and risk: a cross-sectional survey of the impact of COVID-19 on GPs in the first UK city to lock down. BJGP Open. 2020;5(1).

15. Jefferson L, Golder S, Heathcote C, Castro AA, Dale V, Essex H, et al. General practitioner wellbeing during the COVID-19 pandemic: a systematic review. British Journal of General Practice. 2022;In press.

16. GMC. Caring for doctors, caring for patients: how to transform UK healthcare environments to support doctors and medical students to care for patients. Available online: https://www.gmc-uk.org/-/media/documents/caring-for-doctors-caring-for-patients_pdf-80706341.pdf (last accessed 05/08/21). 2019.

17. Deci E, Ryan R. Intrinsic motivation and self-determination in human behavior. US: Springer; 1985.

18. Ritchie J, Spencer L. Qualitative Data Analysis for Applied Policy Research. In: Bryman, Burgess., editors. Analyzing Qualitative Data. London: Routledge; 1994.

19. General Medical Council. The state of medical education and practice in the UK: 2020. Reference tables - Table 19. Available online at: https://www.gmc-ukorg/-/media/documents/gmc-somep-2020-reference-tables-about-the-register-of-medical-practitioners_pdf-84716406pdf (last accessed 17/11/21). 2020.

20. Morgantini LA, Naha U, Wang H, Francavilla S, Acar O, Flores JM, et al. Factors contributing to healthcare professional burnout during the COVID-19 pandemic: A rapid turnaround global survey. PLoS One. 2020;15(9):e0238217.

21. Lange M, Joo S, Couette PA, Le Bas F, Humbert X. Impact on mental health of the COVID-19 outbreak among general practitioners during the sanitary lockdown period. Irish Journal of Medical Science. 2021.

22. Ortega-Galan AM, Ruiz-Fernandez MD, Lirola MJ, Ramos-Pichardo JD, Ibanez-Masero O, Cabrera-Troya J, et al. Professional Quality of Life and Perceived Stress in Health Professionals before COVID-19 in Spain: Primary and Hospital Care. Healthcare. 2020;8(4):10.

23. BMA, RCGP. COVID-19 workload prioritisation unified guidance. London 2021.

24. Stafie CS, Profire L, Apostol MM, Costache II. The professional and psycho-emotional impact of the COVID-19 pandemic on medical care-a Romanian GPS’ perspective. International Journal of Environmental Research and Public Health. 2021;18(4):1–14.

25. Vilovic T, Bozic J, Vilovic M, Rusic D, Furlan SZ, Rada M, et al. Family physicians’ standpoint and mental health assessment in the light of COVID-19 pandemic-a nationwide survey study. International Journal of Environmental Research and Public Health. 2021;18(4):1–17.

26. Baptista S, Teixeira A, Castro L, Cunha M, Serrao C, Rodrigues A, et al. Physician Burnout in Primary Care during the COVID-19 Pandemic: A Cross-Sectional Study in Portugal. Journal of primary care & community health. 2021;32:21501327211008437.

27. Monterrosa-Castro A, Redondo-Mendoza V, Mercado-Lara M. Psychosocial factors associated with symptoms of generalized anxiety disorder in general practitioners during the COVID-19 pandemic. J Investig Med. 2020;68(7):1228–34.

28. Soares A, Thakker P, Deych E, Jain S, Bhayani RK. The Impact of COVID-19 on Dual-Physician Couples: A Disproportionate Burden on Women Physicians. J Womens Health (Larchmt). 2021;30(5):665–71.

29. Adisa TA AO, Adekoya OD. The work–family balance of British working women during the COVID-19 pandemic. Journal of Work-Applied Management. 2021.

30. Martucci S. He’s Working from Home and I’m at Home Trying to Work: Experiences of Childcare and the Work–Family Balance Among Mothers During COVID-19. Journal of Family Issues. 2021.

31. Filfilan NN, Alzhrani EY, Algethamy RF, Fattah RH, Alshehri WA, Althobaity BA, et al. Psychosocial Impact of COVID-19 on Family Physicians in the Kingdom of Saudi Arabia. World Family Medicine. 2020;18(12):91–7.

32. Zeng X, Peng T, Hao X, Zou C, Lin K, Liao X, et al. Psychological Distress Reported by Primary Care Physicians in China during the COVID-19 Pandemic. Psychosomatic Medicine. 2021;83(4):380–6.

33. Pearce C. GPs fear Mail campaign for ‘default’ face-to-face appointments will fuel abuse. Available online at: https://www.pulsetodaycouk/news/breaking-news/gps-fear-mail-campaign-for-default-face-to-face-appointments-will-fuel-abuse/ (last accessed 17/11/21). 2021.

34. NHS England and NHS Improvement. Network Contract Directed Enhanced Service: guidance for 2020/21 in England. Available online: https://www.england.nhs.uk/wp-content/uploads/2020/03/network-contract-des-guidance-2020-21.pdf (last accessed 05/08/21). 2020.

